# Interleukin-6 as a diagnostics marker of intra-amniotic inflammation in PPROM: a systematic review

**DOI:** 10.1101/2025.05.18.25327857

**Authors:** Marie Vajrychova, Michaela Sadibolova, Rudolf Kukla, Radka Bolehovska, Jaroslav Stranik

## Abstract

**Objective:** Intra-amniotic inflammation (IAI) is a frequent complication occurring in preterm prelabour rupture of membrane (PPROM). We report a systematic review to assess the diagnostic potential of interleukin-6 (IL-6) as a marker of IAI.

**Data sources:** For the purpose of this study, we followed a prospective protocol (International Prospective Register of Systematic Reviews, reg. CRD42024501132). We searched PubMed, Web of Science, Scopus, and ResearchRabbit from inception to March 2024.

**Study eligibility criteria:** We included all eligible research articles reporting the concentration of IL-6 in association with IAI in PPROM. Along with amniotic fluid IL-6, prospective and retrospective cohort studies reporting neonatal morbidities, the correlation of IL-6 in amniotic fluid and IL-6 or other proteins in non-invasively collected samples (maternal blood, cervical fluid, and vaginal fluid) were included.

**Methods:** The quality assessment of included studies was performed based on MINORS scoring for non-randomized comparative studies. The extracted data was analyzed using R programming language. Continuous outcomes were analyzed using the median difference and 95% confidence intervals under the inverse variance analysis method (random-effects model). Dichotomous outcomes were reported as risk ratios and 95% confidence intervals.

**Results:** Of 1,133 records screened, we ultimately included 36 eligible studies. The vast majority of studies defined IAI with the threshold of amniotic fluid IL-6 at 2,600 pg/mL for ELISA, 3,000 pg/mL for ECLIA, and 745 pg/mL, if a lateral flow-based immunoassay point-of-care (POC) test was used. IAI was mostly defined according to IL-6 concentration in amniotic fluid, but there was a large-sized positive correlation with IL-6 concentration in cervical fluid and vaginal fluid. Also, IL-6 concentration positively reflected the response of reported proinflammatory proteins in amniotic fluid and cervical fluid (IL-8, MCP-1). Elevated concentration of IL-6 was associated with a higher proportion of bronchopulmonary dysplasia, respiratory distress syndrome, and early-onset neonatal sepsis. Finally, the occurrence of *Ch. trachomatis*, *F. nucleatum*, and *S. anginosus* was more frequent in microbial-associated IAI.

**Conclusion:** Evaluating data from all included studies, we summarized that IL-6 is a versatile and worthwhile diagnostics marker for the diagnosis of microbial-associated and sterile IAI in PPROM with a potential to recognize IAI also in non-invasively collected samples.

**Global Reports at a Glance:** *Why was this study conducted?:* IAI in PPROM is a subclinical pathological state representing a risk of severe consequences for newborns. The question of the diagnostic potential of amniotic fluid IL-6 has been raised in the last two decades since there is a lack of reliable markers easily available in maternal blood. In this study, we therefore summarized the knowledge about IL-6 to assess its ability to uncover the ongoing but hidden inflammatory response associated with short-term neonatal outcome.

*Key findings:* IL-6 is a valuable marker of microbial-associated and sterile IAI easily available in amniotic fluid as well as non-invasively collected cervical and vaginal fluids.

*What does this add to what is known?:* This review demonstrates that IL-6 is a valuable marker of IAI associated with short-term neonatal morbidities in PPROM. Quantification of IL-6 enables the distinction of microbial-associated and sterile IAI and might also contribute to the diagnostics of IAI using non-invasively collected cervical or vaginal fluids.

## Introduction

It is estimated that every year 15 million infants are born prematurely (1), and one million die due to complications of preterm birth (2). Unfortunately, the rate of preterm birth is rising in many countries (3). Approximately 30% of all spontaneous preterm deliveries represent preterm prelabour rupture of membranes (PPROM), a preterm birth phenotype defined as leakage of amniotic fluid without regular uterine activity prior to 37 weeks of gestation (4,5). A large portion of PPROM cases is complicated by the elevation of inflammatory proteins in amniotic fluid, which is defined as intra-amniotic inflammation (IAI) (6,7). IAI can be triggered by microbes ascending from the lower genital tract into the amniotic fluid, which is a situation called microbial-associated IAI (8). IAI can also be sterile when inflammation arises without the proven presence of microbes in the amniotic fluid (7,9). No matter the associated microbial presence, several studies showed that the IAI is linked to worsened pregnancy outcomes, mainly to a higher rate of neonatal complications (7,10). Due to this fact, the identification of pregnancies complicated by IAI is important for clinical management and can be key in efforts to improve pregnancy outcomes of PPROM.

Amniotic fluid obtained by amniocentesis is the most direct biological substance for IAI diagnostics. Several markers, including glucose, matrix metalloproteinase-8, and interleukine-6 (IL-6), have been shown to be effective in diagnosing IAI in amniotic fluid (7,11,12). Nowadays, IL-6 is a frequently used marker for IAI in amniotic fluid, as several diagnostic methods are available for clinical use (13). However, the threshold value of amniotic fluid IL-6 for the identification of IAI varies across the research articles (13–15). It may be dependent on diagnostic methods and also on gestational age.

Amniocentesis to obtain amniotic fluid is an invasive procedure. Due to low amniotic fluid amounts in women with PPROM, amniocentesis is also demanding and not always feasible (16). Several non-invasively or minimally-invasively obtained biological materials, like vaginal and cervical fluids or maternal blood, can potentially reflect the intra-amniotic environment and be used for IAI diagnosis (17). IL-6 with a specific cut-off value, can be useful for identifying PPROM women with IAI, also in the case of non- or minimally-invasively obtained biological materials.

This study aims to review articles that used defined concentrations of IL-6 in amniotic fluid, and also in non-/ minimally-invasively collected maternal blood, cervical and vaginal fluid to recognize IAI in women with PPROM. We aimed to assess the potential of IL-6 with regards to microbial-associated and sterile IAI stratification, the prediction of neonatal morbidity, and the transfer the diagnostics of IAI from amniotic fluid collection to using non-/ minimally-invasively collected samples.

## Methods

### Search strategy

The literature search was performed in accordance with the Preferred Reporting Items for Systematic Reviews and Meta-Analyses (i.e., “PRISMA”) checklist/methodology (18). This systematic review was registered in the PROSPERO database (registration number CRD42024501132). The literature search was performed between January and March 2024 using the following databases: PubMed, Scopus, Web of Science, and ResearchRabbit. The following terms, adjusted for the database, were used to retrieve the articles of interest: “IL-6”, “PPROM”, “interleukin-6”, “intra-amniotic inflammation”.

### Study selection

All studies retrieved in the literature search were screened using the Rayyan app (19). Duplicates were removed, and all remaining studies were screened according to title and abstract by two reviewers (M.V and J.S.). Only those studies that potentially met the inclusion criteria were selected for full-text reading, of which only those that fulfilled all inclusion criteria were included. In case of disagreement, the other authors of the study (M.S., R.K., and R.B.) and the senior and the most experienced author (M.K.) rechecked the studies and made the final decision.

### Criteria for study selection

Criteria for study inclusion were as follows: women older than 18 years with a singleton pregnancy and with the diagnosis of PPROM. Exclusion criteria were pregnancy complications such as intrauterine growth restriction, structural malformation or chromosomal abnormality of the fetus, maternal hypertension, diabetes mellitus, pre-eclampsia, thyroid disease, vaginal bleeding, and fetal hypoxia. Articles that included women with preterm labor with intact membranes or iatrogenic preterm delivery were excluded from this review.

There were no limitations to the year of publication. All study designs, except for letters, comments, reviews, and practice guidelines, were considered to be potentially eligible. Studies involving animal models and in vitro studies using cell cultures, cell lines, and stem cells were excluded.

We restricted the selection to studies written in English.

### Quality assessment

Four investigators (M.V., J.S., M.S., R.K.) independently assessed the risk of bias using the Methodological index for nonrandomized studies (MINORS) checklist (12 items). A 3-point scale was used to grade the quality of each item on the MINORS checklist (0-not reported, 1-reported but inadequate, 2-reported and adequate). In case of any disagreement between the examiners after independent evaluation, consensus was reached by re-evaluation and discussion with a more experienced author (M.K.).

### Data extraction

Extracted dataset contained the first author, title, publication year, journal, analytical method for IL-6 measurement, cut-off value of IL-6 for IAI diagnostics, collected biological material, number of women, gestational age of included women, diagnostics of the presence of microbes and other markers in a biological sample, recorded short term neonatal outcome, correlation of IL-6 levels in amniotic fluid and other biological material. We summarize the main findings about IL-6 cut-off for qualitative analysis. For quantitative analysis, we summarize the main findings about IL-6 concentration (median + IQR) in microbial-associated IAI and sterile IAI, correlation of IL-6 with other proteins and correlation of IL-6 in invasive (amniotic fluid) and non-invasive model (cervical fluid, vaginal fluid, plasma/serum), proportion of neonatal morbidity and the presence of microbial invasion of amniotic cavity (MIAC). Also, all required data for risk of bias assessment was retrieved.

### Statistical analysis

We analyzed data using R programming language in R studio (RStudio: Integrated Development Environment for R. Posit Software, PBC, Boston, MA. URL http://www.posit.co/) using R packages meta (20), metafor (21), dmetar (22), metamedian (23), and tidyverse (24) based on the guide provided by Harrer et al. 2021 (25). The random-effects model was used for data analysis. Continuous outcomes were analyzed using median difference and 95% confidence intervals (CIs) and using correlation coefficients and CIs. Dichotomous outcomes were analyzed using risk ratios (RR) and 95% CIs. The heterogeneity among the studies was assessed using the P value of the chi-square test and the I2 statistic. The outcome was considered heterogeneous if p<.1 or if I2 > 50%.

## Results

### Study selection and characteristics

A total 1461 articles with publication dates from 1981 to 2024 were found through the screened databases. Duplicates (n=328) were removed before screening. We excluded 1019 articles from the following reasons: reviews, non-human studies, other diagnosis than PPROM, other analytical method than ELISA, ECLIA, and POC, abstract and/or full text not available, contents not related to our review, or significant information about IL-6 concentration not provided. A total number of 114 articles were included for full text screening. Finally, we reviewed 36 articles characterized in Table 1 (6,9,11,13,15,16,26–55). The process of study selection is summarized in PRISMA flowchart (Fig. 1). We included 14 prospective (9,16,27,29,30,38–43,47,48,54) and 18 retrospective cohort studies (6,11,13,15,26,31,32,34–37,44–46,49,50,52), 1 cross-sectional study (53), and 1 nested case-control study (28). The mean of cohort size was 145.4 ± 85.7 and all samples were collected from women with the diagnosis of PPROM between 20^th^ and 37^th^ week of gestation. MIAC was diagnosed using amniotic fluid culture in 9 studies, using PCR in 1 study, and using amniotic fluid culture and PCR in 24 studies. The amniotic fluid was collected in 34 included studies, cervical fluid in 7 included studies, vaginal fluid in 2 studies, and maternal blood (or plasma, or serum) in 7 included studies (Table 2).

**Figure 1:**
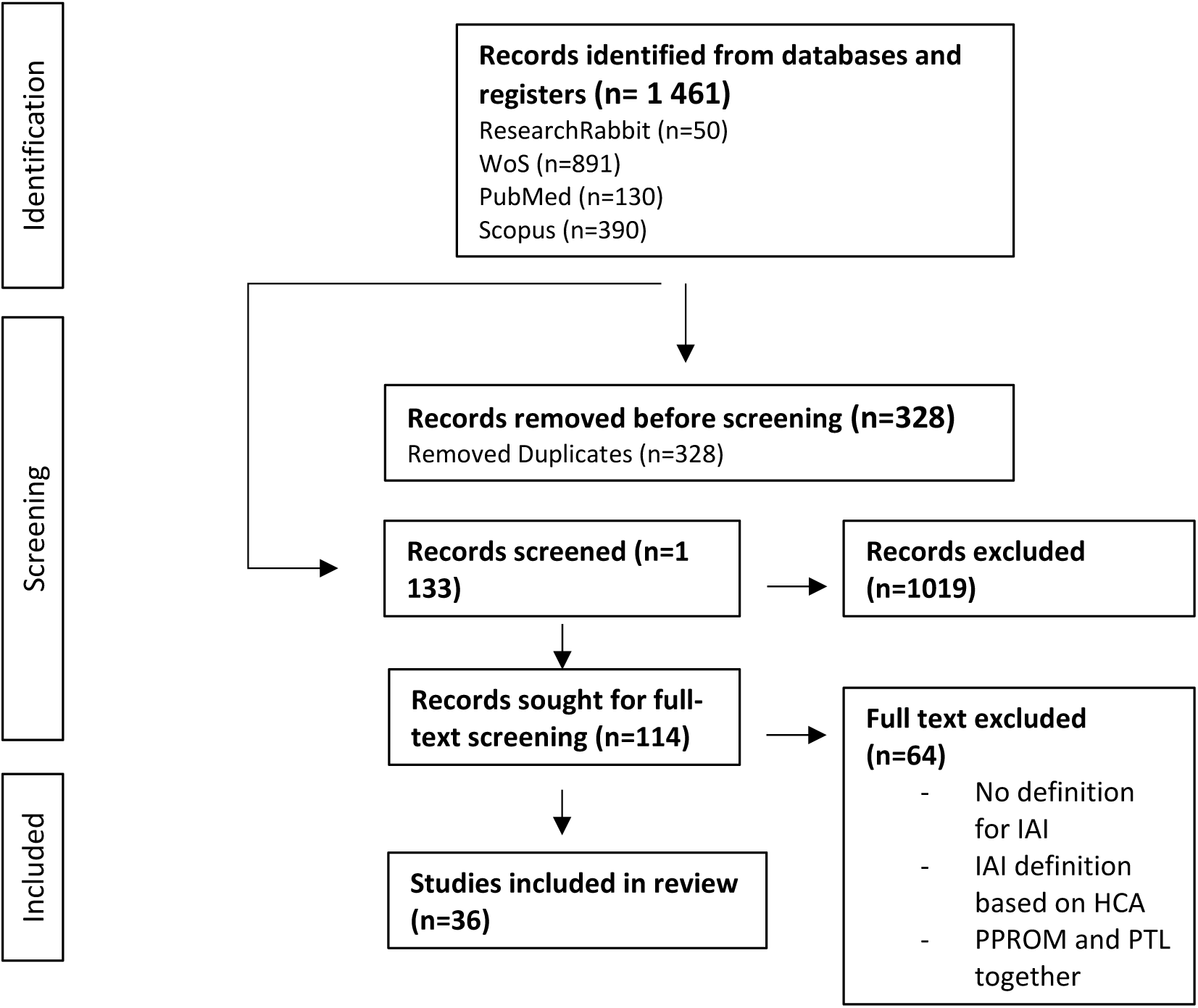
PRISMA flow diagram Abbreviations: PRISMA – Preferred Reporting Items for Systematic Reviews and Meta-Analyses

**Table 1:** Baseline characteristics of the included studies. Abbreviations: GA – gestational age, AF – amniotic fluid, CF – cervical fluid, IAI – intra-amniotic inflammation, POC – lateral flow-based immunoassay point-of-care test

| Studies | Study design | GA | Sample size | Main findings |
| --- | --- | --- | --- | --- |
| Aberšek et al., 2024 (26) | retrospective cohort study | 24+0 - 36+6 | 114 | IAI was defined as an AF IL-6 level $\geq 3000$ pg/mL measured by ECLIA based on previous report. |
| Andrys et al., 2018 (27) | prospective cohort study | 24+0 - 36+6 | 80 | IAI was defined as a concentration of bedside AF IL-6 745 pg/mL based on previous reports. |
| Back et al., 2023 (28) | nested case–control study | 23+0 - 33+6 | 182 | IAI was defined as an AF IL-6 level $\geq 2600$ pg/mL based on previous reports. |
| Chaemsaitong et al., 2016 (15) | retrospective cohort study | 20-35 | 56 | IAI was defined as an AF IL-6 level $\geq 2600$ pg/mL based on previous reports. |
| Fulova et al., 2021 (29) | prospective cohort study | 24+0 - 34+0 | 62 | IAI was defined as AF IL-6 1000 pg/mL (bed-side test). The authors didn't offer an explanation for this specific cut-off. |
| Gulati et al., 2012 (30) | prospective cohort study | 24+0 - 34+0 | 45 | Maternal serum IL-6 levels $\geq 8$ pg/ml for infectious morbidity. |
| Ikeda et al., 2023 (31) | retrospective cohort study | 22+0 - 33+6 | 69 | IAI = AF IL-6 level of $\geq 2600$ pg/mL using CLEIA. A correlation between CLEIA and ECLIA for IL-6 was high ($\rho = 1.00$). Cut-off AF IL-6 = 3000 pg/mL using ECLIA. |
| Joo et al., 2021 (32) | retrospective cohort study | 24+0 - 33+6 | 168 | IAI was defined as an AF IL-6 level $\geq 2600$ pg/mL based on previous reports. |
| Jun et al., 2000 (33) | | N/A | 86 | IAI was defined as CF IL-6 concentration $>350$ pg/mL. |
| Kacerovsky et al., 2018 (16) | prospective cohort study | 24+0 - 36+6 | 118 | The VF IL-6 concentration 2500 pg/mL was tested as cut-off value. |
| Kacerovsky et al., 2020 (34) | retrospective cohort study | 24+0 - 33+6 | 270 | IAI was defined as a concentration of bedside AF IL-6 745 pg/mL based on previous reports. |
| Kacerovsky et al., 2022a (11) | retrospective cohort study | 24+0 - 36+6 | 142 | IAI was defined as an AF IL-6 level $\geq 3000$ pg/mL measured by ECLIA based on previous report. |
| Kacerovsky et al., 2022b (35) | retrospective cohort study | 24+0 - 36+6 | 302 | IAI was defined as an AF IL-6 level $\geq 3000$ pg/mL measured by ECLIA based on previous report. |
| Kacerovsky et al., 2022c (36) | retrospective cohort study | 24+0 - 36+6 | 217 | IAI was defined as a concentration of bedside AF IL-6 745 pg/mL based on previous reports. |
| Lee et al., 2018 (37) | retrospective cohort study | 21+0-34+0 | 75 | Levels of inflammatory proteins measured in the CVF were significantly correlated with AF proteins levels. |
| Lewis et al., 2001 (38) | prospective cohort study | 24+0 - 35+0 | 57 | The diagnosis of IAI was defined as a concentration of ELISA IL-6 in maternal serum of 18 pg/mL. |
| Maymon et al., 2001 (55) | Not defined | <36 | 101 | IAI defined as detection of a positive AF culture in patients with preterm PROM. |
| Musilova et al., 2015 (9) | prospective cohort study | 24+0 - 36+6 | 166 | IAI was defined as a concentration of bedside AF IL-6 745 pg/mL based on previous reports. |
| Musilova et al., 2016a (39) | prospective cohort study | 24+0 - 36+6 | 145 | IAI was defined as a concentration of bedside AF IL-6 745 pg/mL based on previous reports. |
| Musilova et al., 2016b (40) | prospective cohort study | 24+0 - 36+6 | 141 | A cutoff value of vaginal fluid IL-6 of 2500 pg/mL. IAI was defined as a concentration of bedside AF IL-6 745 pg/mL based on previous studies. |
| Musilova et al., 2016c (41) | prospective cohort study | 24+0 - 36+6 | 168 | IAI was defined as a concentration of bedside AF IL-6 745 pg/mL based on previous reports. |
| Musilova et al., 2017a (44) | retrospective cohort study | 24+0 - 36+6 | 136 | IAI was defined as a concentration of bedside AF IL-6 745 pg/mL based on previous reports. |
| Musilova et al., 2017b (45) | retrospective cohort study | 24+0 - 36+6 | 154 | IAI was defined as a concentration of bedside AF IL-6 745 pg/mL based on previous reports. |
| Musilova et al., 2017c (42) | prospective cohort study | 24+0 - 36+6 | 479 | IAI was defined as a concentration of bedside AF IL-6 745 pg/mL based on previous reports. |
| Musilova et al., 2017d (43) | prospective cohort study | 24+0 - 36+6 | 287 | IAI was defined as a concentration of bedside AF IL-6 745 pg/mL based on previous reports. |
| Musilova et al., 2018a (13) | retrospective cohort study | 24+0 - 36+6 | 120 | IL-6 concentrations were assessed using both the ECLIA (3000 pg/mL) and ELISA. A correlation between both assays was found (Spearman's rho = 0.97; p<.0001). |
| Musilova et al., 2018b (54) | prospective cohort study | 24+0 - 36+6 | 144 | IAI was defined as a concentration of bedside AF IL-6 745 pg/mL based on previous reports. |
| Park et al., 2012 (46) | retrospective cohort study | 20+0 - 34+6 | 171 | The best cutoff value for AF IL-6 in predicting the presence of intra-amniotic infection was 2.6 ng/mL and was thus used to diagnose the presence of IAI. |
| Rizzo et al., 1998 (47) | prospective cohort study | 24+0 - 32+0 | 124 | Cut-off IL-6 level in CF 1200 pg/mL. Cut-off AF IL-6 10000 pg/mL. Both determined based on positive AF culture. |
| Rodríguez-Trujillo et al., 2016 (48) | prospective cohort study | 22+0–34+0 | 165 | IAI was defined as a concentration of AF IL-6 1430 pg/mL based on previous reports. |
| Romero et al., 2015 (6) | retrospective cohort study | 20-35 | 59 | IAI was defined as an AF IL-6 level ≥ 2600 pg/mL (ELISA) based on previous reports. |
| Soucek et al., 2023 (49) | retrospective cohort study | 24+0 - 36+6 | 80 | IAI was defined as an AF IL-6 level ≥ 3000 pg/mL measured by ECLIA based on previous report. |
| Spacek et al., 2022a (50) | retrospective cohort study | 24+0 - 36+6 | 144 | IAI was defined as an AF IL-6 level ≥ 3000 pg/mL measured by ECLIA based on previous report. |
| Spacek et al., 2022b (51) | retrospective cohort study | 24+0 - 36+6 | 166 | IAI was defined as an AF IL-6 level $\geq 3000$ pg/mL measured by ECLIA based on previous report. |
| Stranik et al., 2021 (52) | retrospective cohort study | 24+0 - 36+6 | 170 | IAI was defined as an AF IL-6 level $\geq 3000$ pg/mL measured by ECLIA based on previous report. |
| Theis et al., 2020 (53) | retrospective cross-sectional study | 22+0 - 34+0 | 59 | IAI was defined as an AF IL-6 level $\geq 2600$ pg/mL (ELISA) based on previous reports. |

**Table 2:** IL-6 cut-off values in the included studies. Abbreviations: CI – confidence interval, AF – amniotic fluid, CF – cervical fluid, MB – maternal blood, MP – maternal plasma, MS – maternal serum, POC – lateral flow-based immunoassay point-of-care test, VF – vaginal fluid

| Studies | Collected samples | Analytical method | IAI definition | Determined cut-off value | Other marker of IAI | Neonatal morbidity | sterile IAI |
| --- | --- | --- | --- | --- | --- | --- | --- |
| Jun et al., 2000 (33) | AF, CF | ELISA | positive AF culture | 350 (CF) |  | yes | no |
| Lee et al., 2018 (37) | AF, CF, MP | multiplex immunoassay | positive AF culture | 1488.7 (AF), 310.79 (CF) | IL-8, MCP-1, MIP-1alpha, MIP-2beta | no | no |
| Musilova et al., 2016b (40) | AF, VF | POC | AF IL-6 (cut-off 745 pg/mL) | 2500 (VF) |  | no | yes |
| Musilova et al., 2018a (13) | AF | ECLIA | AF IL-6 (cut-off 2600 pg/mL) | 3000 (AF) |  | no | no |
| Musilova et al., 2018b (54) | AF, CF | POC, ELISA | AF IL-6 (cut-off 745 pg/mL) | 500 (CF) |  | no | yes |
| Rizzo et al., 1998 (47) | AF, CF | ELISA | positive AF culture | 10000 (AF), 200 (CF) |  | no | no |
| Studies | Collected samples | Analytical method | IAI definition | Used IL-6 cut-off | Other marker of IAI | Neonatal morbidity | sterile IAI |
| Aberšek et al., 2024 (26) | AF | ECLIA | IL-6 cut-off | 3000 | IP-10 | no | yes |
| Andrys et al., 2018 (27) | AF, CF | POC | IL-6 cut-off | 745 | CALR, CTSG | no | no |
| Back et al., 2023 (28) | AF, MP | ELISA | IL-6 cut-off | 2600 | FCGR3A, HP, LRP1 | no | yes |
| Chaemsaitong et al., 2016 (15) | AF | ELISA, POC | IL-6 cut-off | 2600, 745 |  | no | no |
| Fulova et al., 2021 (29) | AF | POC (point-of-care) | IL-6 cut-off | 1000 | CRP | yes | yes |
| Gulati et al., 2012 (30) | MS | ELISA | IL-6 cut-off | 8 | CRP | yes | no |
| Ikeda et al., 2023 (31) | AF | CLEIA/ECLIA | IL-6 cut-off | 3000 |  | yes | no |
| Joo et al., 2021 (32) | AF, MP | ELISA | IL-6 cut-off | 2600 | IL-8, CRP, LBP, PTX3, RETN, IGFBP-3 |  | yes |
| Kacerovsky et al., 2018 (16) | AF, VF | POC | IL-6 cut-off | 745, 2500 |  | no | yes |
| Kacerovsky et al., 2020 (34) | AF | POC | IL-6 cut-off | 745 |  |  | yes |
| Kacerovsky et al., 2022a (11) | AF | ECLIA | IL-6 cut-off | 3000 | glucose | no | yes |
| Kacerovsky et al., 2022b (35) | AF | ECLIA | IL-6 cut-off | 3000 |  | yes | yes |
| Kacerovsky et al., 2022c (36) | AF, CF | POC, ECLIA | IL-6 cut-off | 745, 3000 |  | no | yes |
| Lewis et al., 2001 (38) | MP | ELISA | not defined |  |  | yes | no |
| Maymon et al., 2001 (55) | AF | ELISA | not defined |  | MMP8 | yes | no |
| Musilova et al., 2015 (9) | AF | POC | IL-6 cut-off | 745 | CRP | yes | yes |
| Musilova et al., 2016a (39) | AF | POC | IL-6 cut-off | 745 | PGE2 | no | yes |
| Musilova et al., 2016c (41) | AF | POC | IL-6 cut-off | 745 | CALR | no | yes |
| Musilova et al., 2017a (44) | AF | POC | IL-6 cut-off | 745 | CLU | no | yes |
| Musilova et al., 2017b (45) | AF | POC | IL-6 cut-off | 745 | CTSG | no | yes |
| Musilova et al., 2017c (42) | AF, MB | POC | IL-6 cut-off | 745 | WBC | no | yes |
| Musilova et al., 2017d (43) | AF, MB | POC | IL-6 cut-off | 745 | CRP | no | yes |
| Park et al., 2012 (46) | AF, MP | ELISA | IL-6 cut-off | 2600 | CRP |  | yes |
| Rodríguez-Trujillo et al., 2016 (48) | AF | ELISA | IL-6 cut-off | 1430 |  | yes | yes |
| Romero et al., 2015 (6) | AF | ELISA | IL-6 cut-off | 2600 |  | no | yes |
| Soucek et al., 2023 (49) | AF | ECLIA | IL-6 cut-off | 3000 | CD36 | no | yes |
| Spacek et al., 2022a (50) | AF | ECLIA | IL-6 cut-off | 3000 | sCD93 | no | yes |
| Spacek et al., 2022b (51) | AF | POC | IL-6 cut-off | 745 | GZMA | no | yes |
| Stranik et al., 2021 (52) | AF, CF | ECLIA | IL-6 cut-off | 3000 | FCGBP | no | yes |
| Theis et al., 2020 (53) | AF | ELISA | IL-6 cut-off | 2600 |  | no | yes |

### Quality assessment

MINORS scores are shown in Figure 2. The median of MINORS score was 22 (range 16 – 24). All included studies were deemed at low risk of bias. However, only one study reported prospective calculation of the study size.

**Figure 2:**
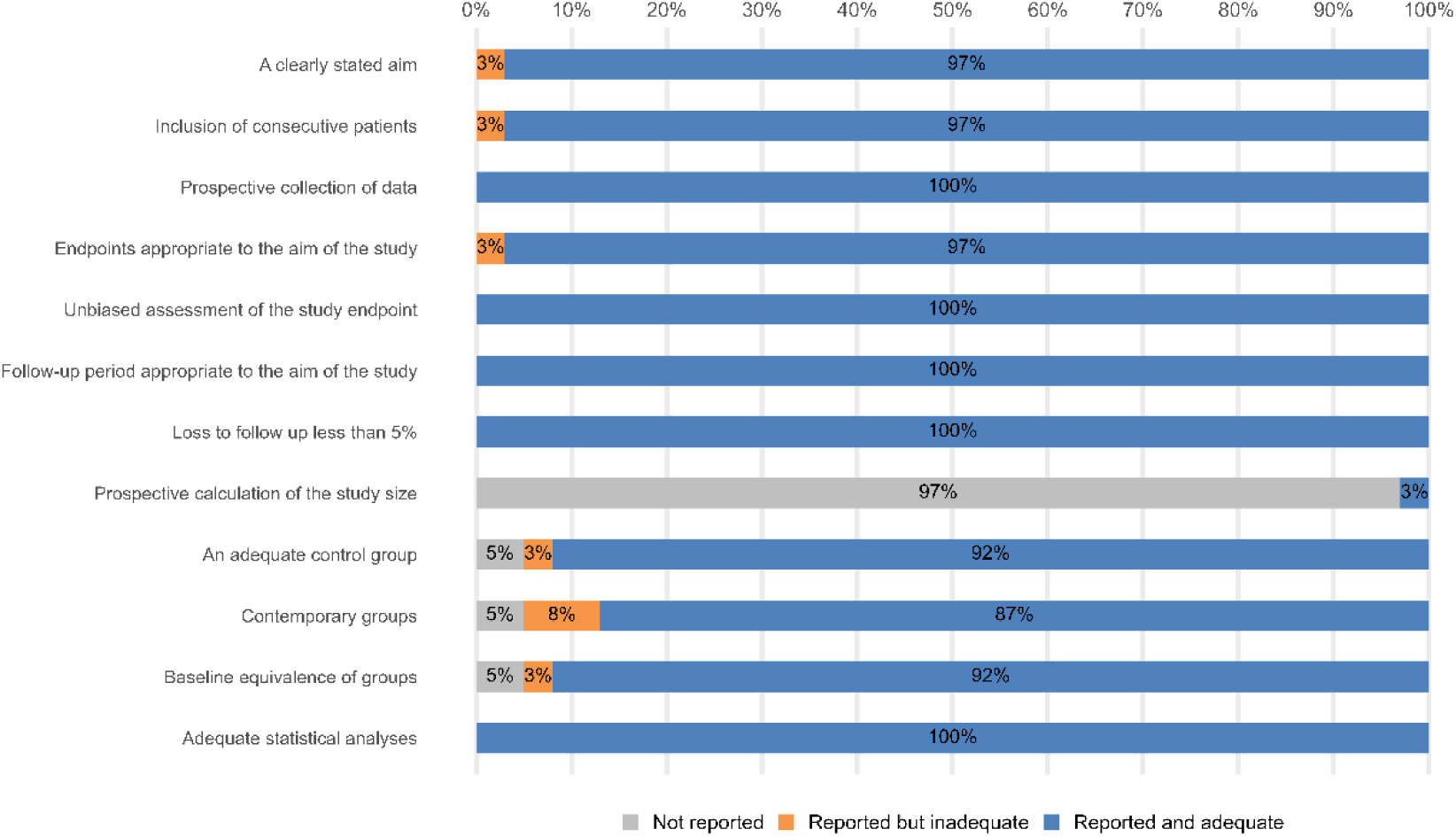
Results of the quality assessment based on the MINORS scoring Abbreviations: MINORS - Methodological index for nonrandomized studies

### Cut-off values for IAI diagnosis

Cut-off values reported in all included studies are summarized in Table 2. Six of all included studies defined new IL-6 cut-off values as the output (13,33,37,40,47,54). Of these, three studies defined IAI based on previously determined amniotic fluid IL-6 cut-off values of 2,600 pg/mL (13) and 745 pg/mL (40,54). Three studies stratified PPROM cohort according to the presence of MIAC (33,37,47). Specificity, sensitivity, positive and negative predictive values, and likelihood ratios reported in these studies are summarized in Table 3. Two studies reported risk ratio (CI 95%) instead of likelihood ratio and one study reported odds ratio (CI 95%). Of these, one study defined amniotic fluid IL-6 concentration of 10,000 pg/mL as cut-off value for ELISA (47) and the second study defined amniotic fluid IL-6 concentration of 3,000 pg/mL as cut-off value for ECLIA (13). Four studies defined four different cervical fluid IL-6 cut-off values of 200 pg/mL (47), 310 pg/mL (37), 350 pg/mL (33), and 500 pg/mL (54). Finally, one study determined IL-6 concentration of 2,500 pg/mL as a cut-off value to diagnose microbial-associated IAI and sterile IAI in vaginal fluid (40). Of all included studies, 28 used cut-off values from different studies (Table 2) and two studies did not define any IL-6 concentration as a cut-off value (38,55). Six studies used IL-6 concentration of 2,600 pg/mL to recognize IAI and determined amniotic fluid IL-6 concentration using ELISA (6,15,28,32,46,53). Seven studies defined amniotic fluid IL-6 concentration and stratified the PPROM subgroups according to IL-6 cut-off value of 3,000 pg/mL using ECLIA (11,26,31,35,49,50,52). Thirteen studies stratified the PPROM subgroups according to amniotic fluid IL-6 cut-off value of 745 pg/mL (15) using POC (9,15,16,27,34,36,39,41–45,51).

**Table 3:** Accuracy of the determined IL-6 cut-off values in the included studies. Abbreviations: CI – confidence interval, AF – amniotic fluid, CF – cervical fluid, VF – vaginal fluid, RR – risk ratio, OR – odds ratio, AUC – area under the curve

| Studies |  | Determined cut-off value | Sensitivity (%) | Specificity (%) | (+) predictive value (%) | (-) predictive value (%) | Likelihood ratio | Material | RR | OR | CI 95% | CI 95% | AUC |
| --- | --- | --- | --- | --- | --- | --- | --- | --- | --- | --- | --- | --- | --- |
| Jun et al., 2000 (33) |  | 350 | 92 | 78 | 41 | 98 |  | CF | 3.2 |  | 1.6 | 6.5 | 0.84 |
| Lee et al., 2018 (37) |  | 1488.7 |  |  |  |  |  | AF |  | 7.9 | 1.82 | 48.707 | 0.87 |
|  |  | 310.79 |  |  |  |  |  | CF |  | 9.44 | 2.75 | 32.425 | 0.81 |
| Musilova et al., 2016b (40) | Microbial-associated IAI | 2500 | 53 | 89 | 63 | 85 | 5 | VF |  |  |  |  |  |
|  | Sterile IAI | 2500 | 74 | 91 | 67 | 94 | 8.4 | VF |  |  |  |  |  |
| Musilova et al., 2018a (13) |  | 3000 | 88 | 99 | 97 | 96 | 76 | AF |  |  |  |  |  |
| Musilova et al., 2018b (54) |  | 500 | 75 | 85 | 42 | 96 | 4.8 | CF |  |  |  |  |  |
| Rizzo et al., 1998 (47) |  | 10000 | 80.9 | 76.8 | 64.1 | 88.7 |  | AF | 5.69 |  | 2.87 | 11.27 | 0.77 |
|  |  | 200 | 78.5 | 73.1 | 60 | 86.9 |  | CF | 4.6 |  | 2.41 | 8.77 | 0.76 |

### Elevation of IL-6 in microbial-associated and sterile IAI

Twenty-six of all included studies stratified IAI according to the IL-6 concentration above the defined cut-off value and the presence of MIAC to microbial-associated and sterile IAI (Table 2). In these studies, the overall frequency of sterile IAI was approximately half that of the frequency of microbial-associated IAI (RR = 0.49, CI 95% [0.4111; 0.5884], p< 0.0001) (Fig. 3). Data were heterogeneous (*I^2^*= 39.1%, p=0.023). Ten included studies reported the IL-6 concentration for these conditions (6,9,34–36,40,42,45,53,54). The concentration of IL-6 in the presence of microbial-associated IAI was found to be significantly increased compared to sterile IAI (Fig. 4). Pooled estimate of the difference of median was 8,357.4 (CI 95%: 5,100.5 – 11,614.3, p<0.0001). Data were heterogeneous (*I^2^*= 95.5%, p < 0.0001).

**Figure 3:**
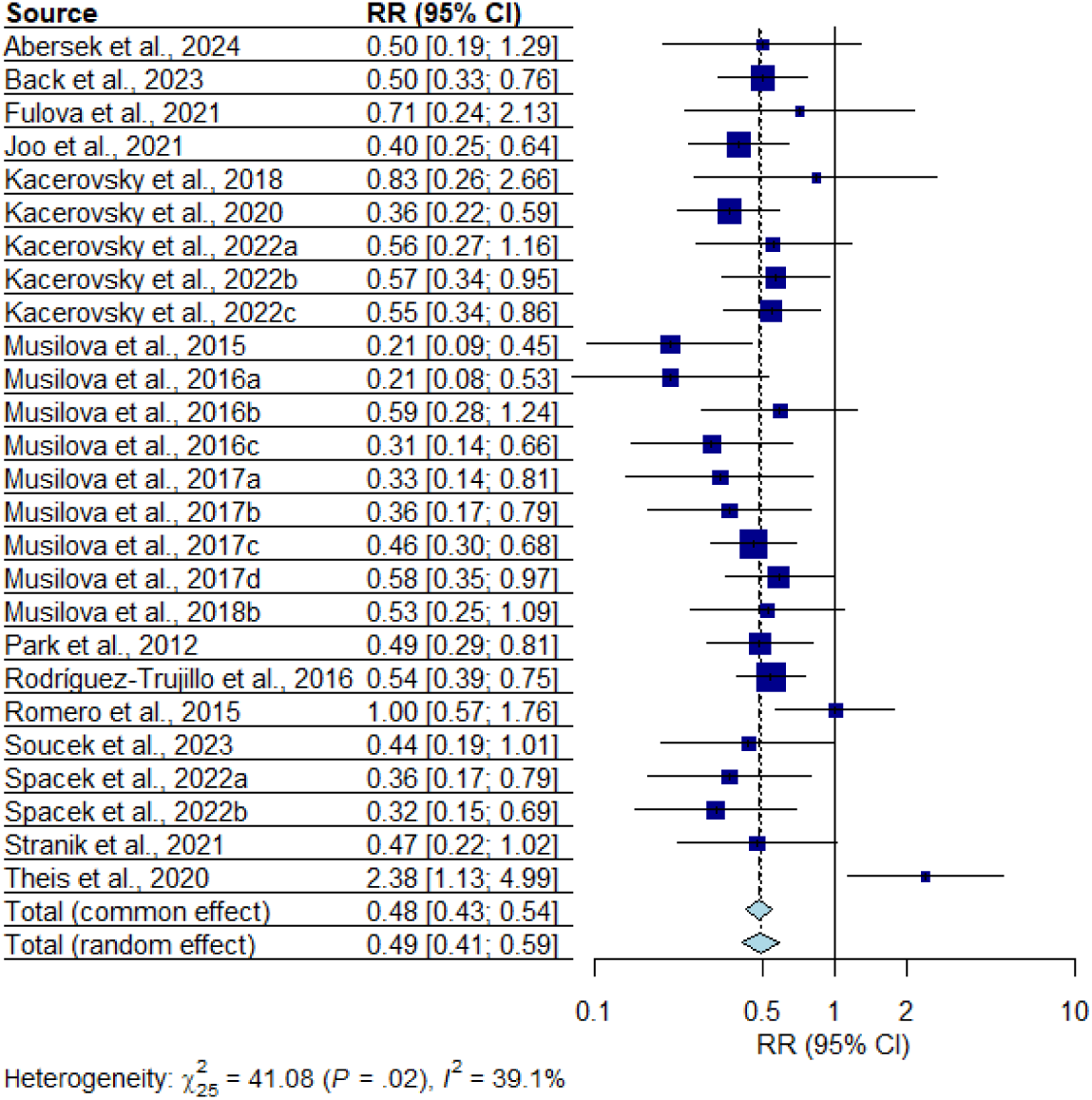
Meta-analysis of the proportion of sterile IAI in PPROM compared to microbial-associated IAI. Abbreviations: CI – confidence interval, RR – risk ratio

**Figure 4:**
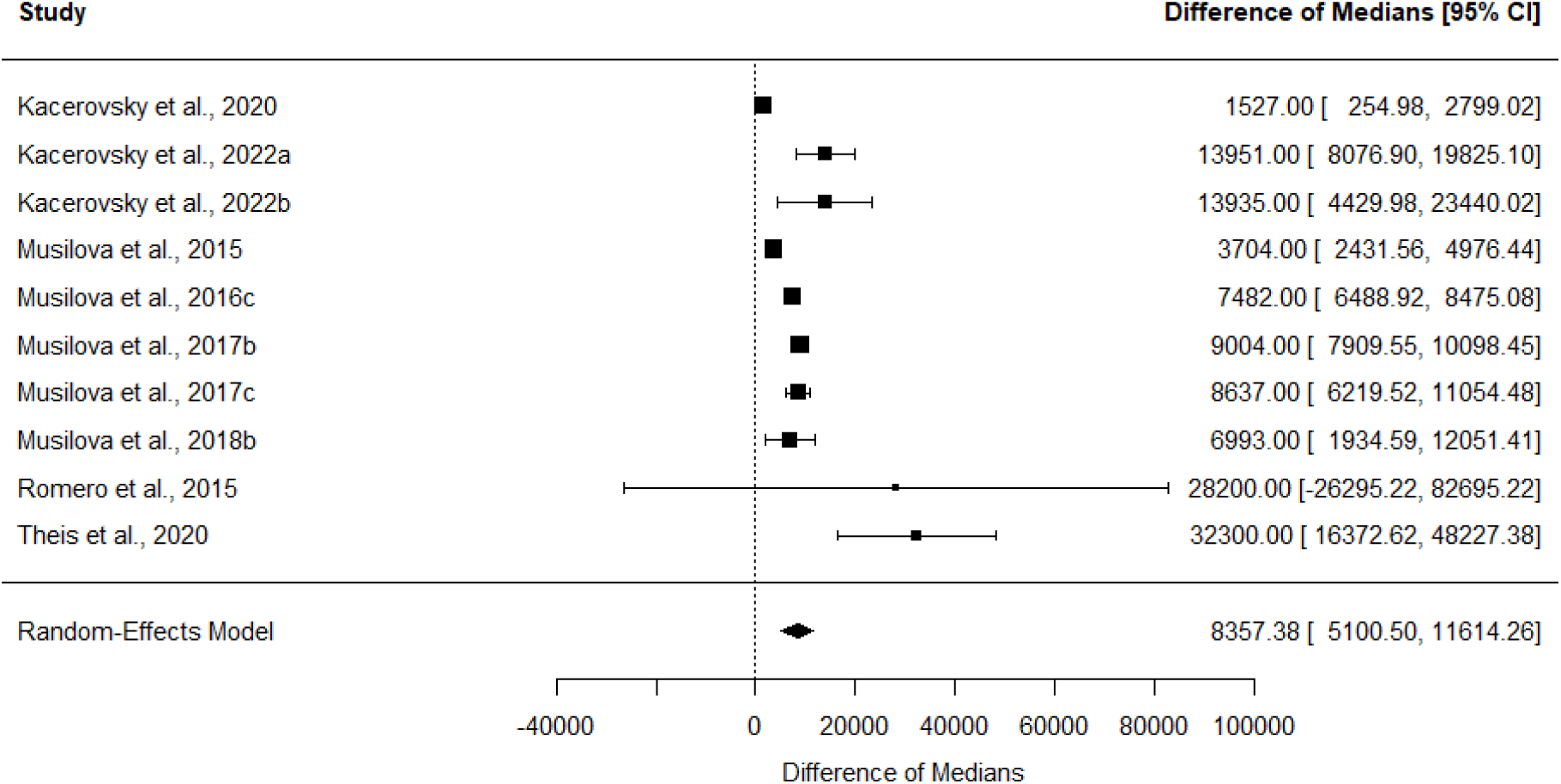
Meta-analysis of IL-6 concentration in microbial-associated IAI and sterile IAI Abbreviations: CI – confidence interval

### Reflection of global proinflammatory response

Of all included studies, 21 studies investigated the level of other potential markers of IAI (selected proteins, prostaglandin E2 (PGE2), WBC, and glucose) in amniotic fluid, cervical fluid, maternal blood, plasma and serum (Table 2) (11,26–30,32,37,39,41–46,49–52,55,56). In these studies, IAI was defined based on the cut-off value of amniotic fluid IL-6. Of these, 11 studies directly determined the correlation between the level of the investigated marker and the IL-6 concentration in amniotic fluid (11,27,29,37,39,41–43,45,49,51). We found a global moderate-sized correlation between IL-6 concentration and the level of other potential IAI markers (0.36, CI 95% [0.126 - 0.561], p = 0.0054) (Fig. 5). Data were heterogeneous (*I^2^* = 95.8%, p < 0.0001).

**Figure 5:**
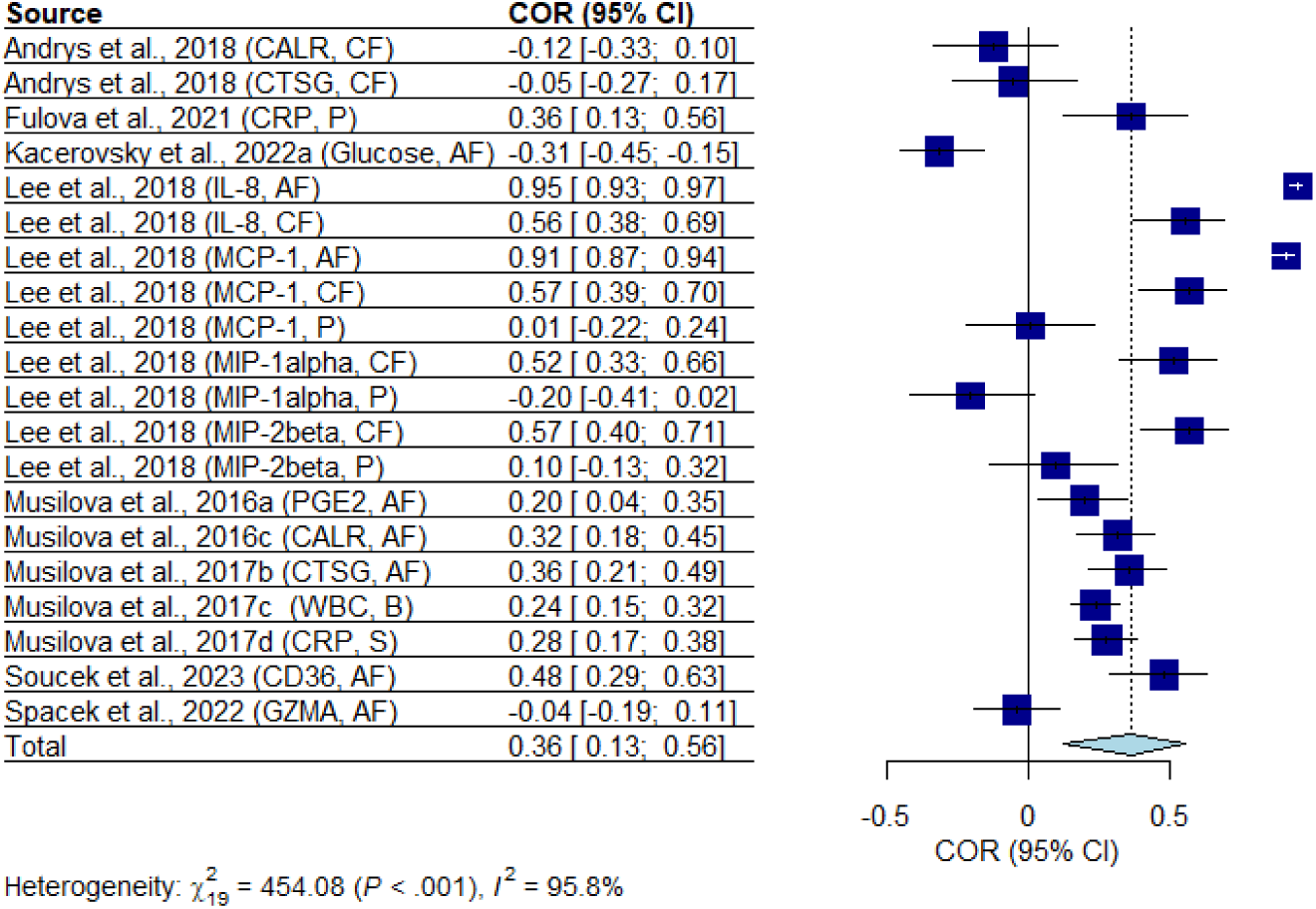
Meta-analysis of the correlation between amniotic fluid IL-6 concentration and the levels of potential protein and non-protein markers of IAI Abbreviations: CI – confidence interval, COR – correlation

### Non-invasive model for diagnosis IAI

A large-sized positive correlation (0.62, CI 95% [0.35 - 0.79], p = 0.0078) was found in 4 studies reporting IL-6 concentrations in amniotic fluid, and cervical fluid, and amniotic fluid and vaginal fluid in parallel (Fig. 6) (33,37,40,47). Data were heterogenous (*I^2^* = 79.1%, p = 0.0025).

**Figure 6:**
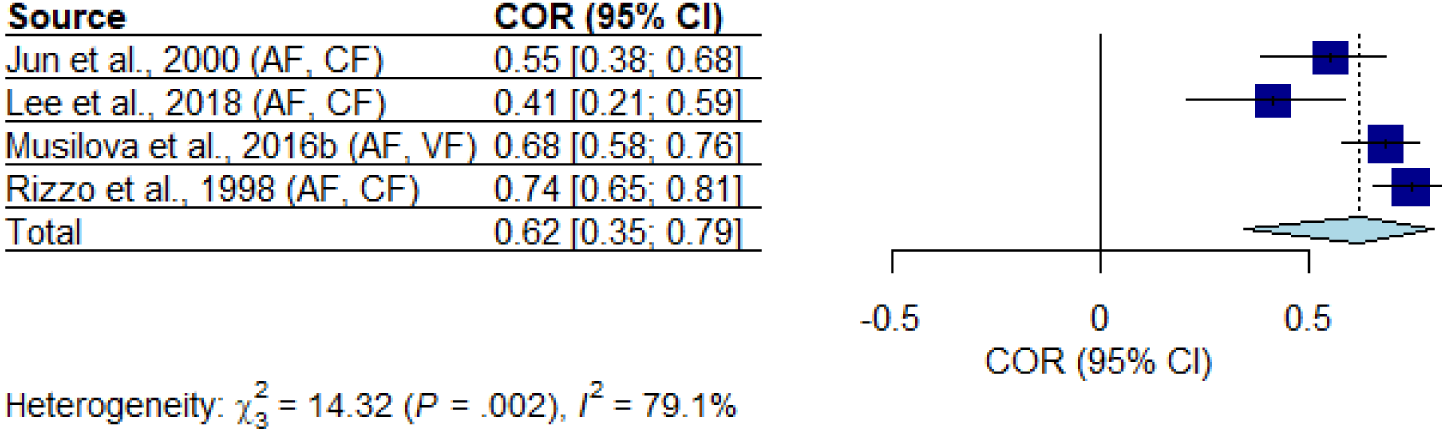
Meta-analysis of the correlation between IL-6 concentrations in amniotic fluid and cervical and vaginal fluid Abbreviations: CI – confidence interval, COR – correlation

### Elevated IL-6 concentration and neonatal morbidity

Of all included studies, nine reported a proportion of neonatal morbidities (29–31,33,35,38,48,55,56). The risk ratio of bronchopulmonary dysplasia (BPD) (RR = 6.14, CI 95% [2.18 - 17.29], p = 0.0082), respiratory distress syndrome (RDS) (RR = 1.72, CI 95% [1.23 - 2.41], p= 0.0075), and early-onset neonatal sepsis (EONS) (RR = 2.51, CI 95% [1.17 - 5.36], p= 0.028) were found significantly higher in the presence of IAI across the included studies in this review. Data were homogenous (BPD: I^2^ = 8.9%, p = 0.356; RDS: I^2^ = 21.4%, p = 0.267; EONS: I^2^ = 0.0%, p = 0.58) (Fig. 7). In addition, four studies divided IAI into microbial-associated IAI and sterile IAI. However, no significant difference in the proportion of neonatal morbidities described above was found when microbial-associated IAI and sterile IAI were compared.

**Figure 7:**
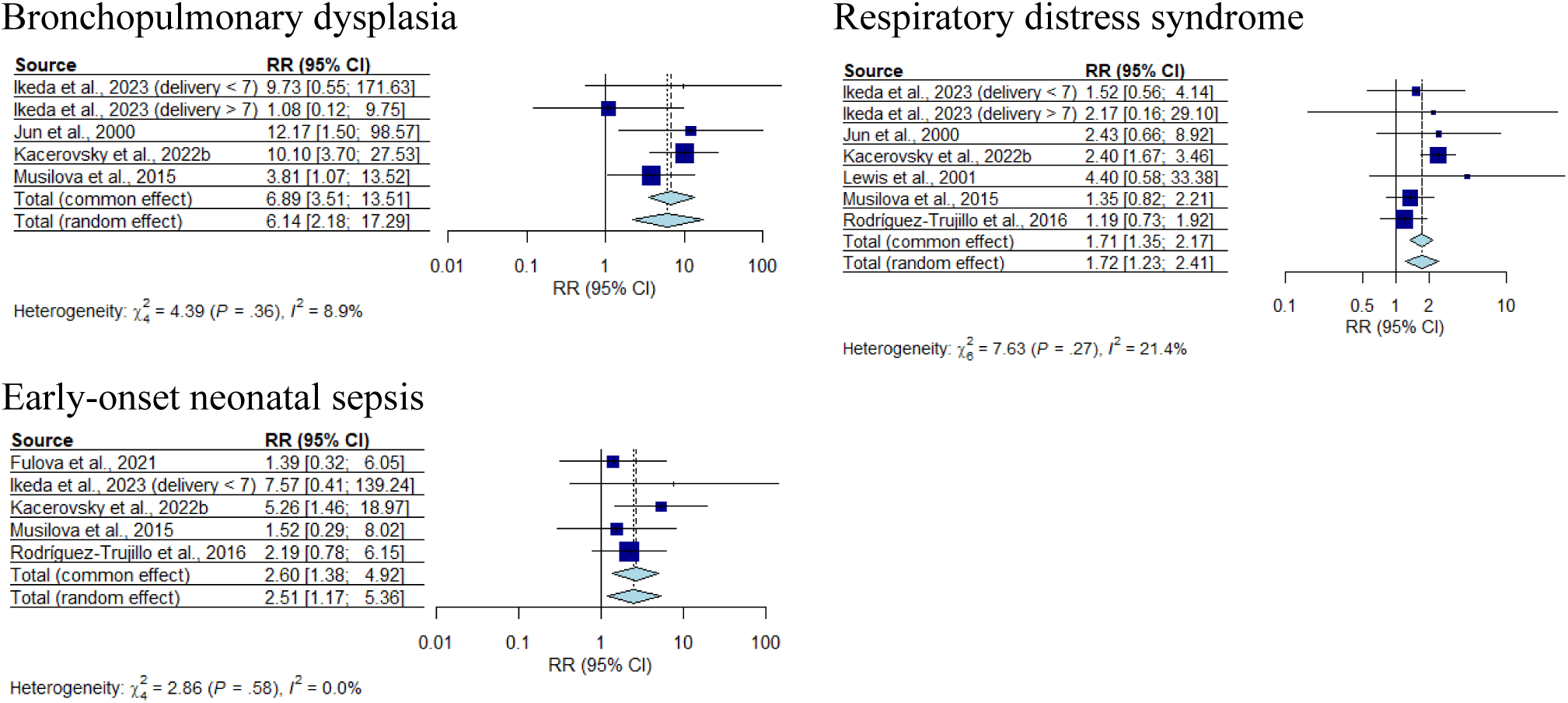
Meta-analysis of the proportion of bronchopulmonary dysplasia, respiratory distress syndrome (RDS), and early-onset sepsis (EONS) in the cases of elevated IL-6 concentration compared to IL-6 concentrations under the defined cut-off values Abbreviations: CI – confidence interval, RR – risk ratio

Elevated IL-6 concentration in the presence of genital mycoplasma and obligatory pathogens Of all included studies, thirteen reported the proportion of bacteria in both microbial-associated IAI and MIAC alone (6,11,31,34–36,41–43,45,51,54,56). Top ten of the most frequent bacteria were investigated in this review (*U. urealyticum, M. hominis, Ch. trachomatis, S. sanquinegens, H. influenzae, G. vaginalis, Streptococcus agalactiae, S. anginosus group, F. nucleatum,* and *S. pneumoniae*). *U. urealyticum* was the most frequent bacterium found in amniotic fluid, but no difference in the proportion was observed in microbial-associated IAI compared to MIAC alone (RR = 0.97, CI 95% [0.87 - 1.08], p = 0.52). Data were homogenous (I^2^ = 23.6%, p = 0.21). On the other hand, we found that the occurrence of *Ch. trachomatis* (RR = 1.5, CI 95% [1.03 - 2.16], p = 0.0356), *S. anginosus* group (RR = 2.60, CI 95% [1.52 - 4.45], p = 0.0029), and *F. nucleatum* (RR = 2.96, CI 95% [2.27 - 3.86], p < 0.0001) was higher in microbial-associated IAI compared to MIAC alone. Data were homogenous (*Ch. trachomatis:* I^2^ = 0%, p = 0.96, *S. anginosus:* I^2^ = 0%, p = 0.99, and *F. nucleatum:* I^2^ = 0%, p = 1.00) (Fig. 8).

**Figure 8:**
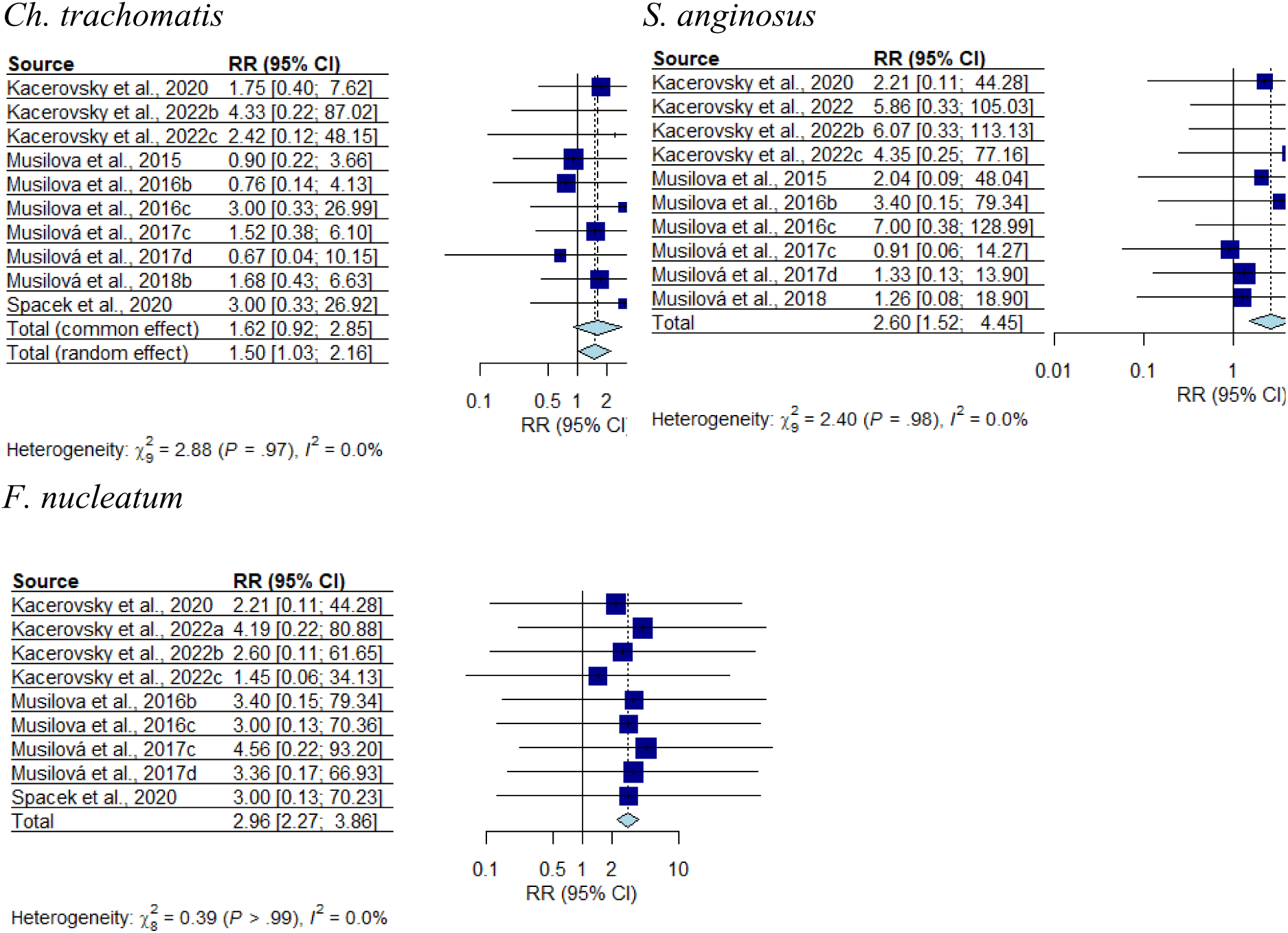
Meta-analysis of the proportion of obligatory pathogens *Ch. trachomatis*, *F. nucleatum*, and *S.anginosus* in the presence of microbial-associated IAI compared to MIAC alone Abbreviations: CI – confidence interval, RR – risk ratio

## Comments

### Principal findings

In this review, we summarized the current knowledge about the previously published cut-off values of IL-6 concentrations for the diagnosis of IAI based on the analytical method and the kind of collected samples. Also, we extracted data about the IL-6 concentration in microbial-associated and sterile IAI, correlation of amniotic fluid IL-6 concentration with other potential protein and non-protein markers of IAI, and correlation with IL-6 concentrations in cervical and vaginal fluid, the proportion of neonatal morbidities and the presence of obligate pathogens along with IAI. The principal findings are as follows: 1) The most frequently used cut-off values for the diagnosis of IAI based on IL-6 concentration in amniotic fluid are 745 pg/mL (POC), 2,600 pg/mL (ELISA), and 3,000 pg/mL (ECLIA). 2) The concentration of amniotic fluid IL-6 is significantly increased in microbial-associated IAI compared to sterile IAI. 3) The levels of other amniotic fluid and cervical fluid proteins, but not plasma or serum proteins, except plasma CRP, moderately correlate with amniotic fluid IL-6 levels as a response to IAI. 4) The concentrations of IL-6 in cervical and vaginal fluid positively correlated with the concentration of amniotic fluid IL-6. 5) The upregulation of amniotic fluid IL-6 is associated with the higher occurrence of bronchopulmonary dysplasia, RDS, and EONS. 6) Increased amniotic fluid IL-6 concentration is significantly associated with the presence of obligate pathogens *Ch. trachomatis*, *S. anginosus*, and *F. nucleatum*.

### Comparison with existing literature

In 1993, Romero et al. determined the level of amniotic fluid IL-6, glucose, WBC, and Gram stain in a cohort of 120 women with preterm labor and intact membranes (PTL) (14). The authors assessed the diagnostic and prognostic value of these tests to recognize MIAC along with short amniocentesis-to-delivery interval and neonatal morbidity. They evaluated that the amniotic fluid IL-6 has the best predictive value for MIAC associated with worsened neonatal outcome and determined the cut-off IL-6 concentration to be 11,300 pg/mL (14). This is in concordance with one study included in this review. Rizzo et al. also reported the amniotic fluid IL-6 concentration of 10,000 pg/mL based on a positive amniotic fluid culture (47). In 2001, Yoon et al. defined a new cut-off value of amniotic fluid IL-6 concentration of 2,600 pg/mL for the diagnosis of IAI in PTL women (57). A new cut-off value was derived from the receiver operating characteristic curve that described the performance of amniotic fluid IL-6 concentration in the identification of a positive amniotic fluid culture. In this study, the portion of women with IL-6 above the cut-off and negative amniotic fluid culture (defined as sterile IAI) had shorter amniocentesis-to-delivery interval, higher frequency of spontaneous preterm delivery, and higher rate of adverse outcome (histological chorioamnionitis, funisitis, early preterm birth, and neonatal morbidity) compared to negative cases. Interestingly, the neonatal outcome did not differ for patients with sterile IAI and patients with microbial-associated IAI (57). The majority of articles included in this review used the cut-off defined by Yoon et al. (57) or a cut-off that was adjusted for the diagnostic method. Several issues can be identified with this approach. a) Yoon et al. in their original research included only women with PTL. PTL and PPROM are two distinct clinical phenotypes of preterm birth with different underlying mechanisms and clinical implications. Both conditions involve inflammation, but the inflammatory profiles differ (58,59). PPROM is characterized by a more pronounced inflammatory response, with higher plasma levels of certain cytokines and chemokines like IL-6 and CXCL1, compared to PTL (59,60). One cut-off may not reflect the same situation for both PTL and PPROM. (b) The cut-off by Yoon et al. (57) was established based on positive amniotic fluid culture. However, the presence of several difficult-to-cultivate microbes associated with MIAC may be proven only by non-cultivation techniques (61,62). Moreover, the same cut-off derived from cases with positive culture is applied to cases with sterile IAI. (c) Physiological levels of IL-6 in amniotic fluid are dynamic and may vary with gestational age (63,64). One cut-off may not be useful for identifying IAI at any gestational age. On the other hand, the Yoon et al. cut-off was tested many times and is connected to worsened maternal and neonatal outcomes even among women with PPROM (15,57). Also, one well-defined cut-off may be useful from a clinical point of view, as it is simple to navigate clinical decisions.

Microbial-associated IAI was related to higher amniotic fluid levels of IL-6 in our study. This is in line with previous literature (6). The microbial-associated IAI is driven, according to the most common theory, by ascending microbes from the lower genital tract (65,66). Microbial presence within the amniotic cavity triggers a massive local immune response that involves both the maternal and the fetal compartments and is characterized by the abundant presence of neutrophils, monocytes/macrophages, and T cells, together with the elevation of inflammatory mediators (such as IL-6) (65).

Sterile IAI in women with PPROM may be attributable to the fetal membrane damage resulting in the release of sterile endogenous molecules (alarmins) into the amniotic fluid, which subsequently triggers an inflammatory response through pattern recognition receptors (65). It can also be caused by the infection in the choriodecidual space, leading to the release of inflammatory mediators from the fetal membranes into the amniotic fluid (7). The frequency of sterile IAI among women with PPROM complicated by IAI in our review is approximately 50% (Fig. 3), which is in concordance with the studies by Shim et al. (7) and Romero et al. (6). However, Musilova et al. showed a lower frequency of sterile IAI (4% sterile IAI vs 21% intra-amniotic infection) (56). The frequency of sterile IAI can be influenced by the gestational age of women in the included cohort. The microbial-associated IAI is more common among women below 25 weeks of gestation, and the frequency of sterile IAI increases with gestational age (6,56).

Identifying non-invasive biomarkers for IAI in women with PPROM holds significant clinical value, especially given the challenges associated with obtaining amniotic fluid through invasive procedures, a particular concern for those often presenting with oligohydramnios (67,68). Non-invasive diagnostic methods using cervical and vaginal fluids show promise in identifying IAI in PPROM cases. Our study showed that there is a positive correlation between amniotic fluid IL-6, identifying IAI, and cervical and vaginal fluid IL-6 and other biomarkers. Chang et al. conducted a systematic review with meta-analysis of nine studies involving 1904 preterm birth patients (mostly PTL) (69). In those, the concentration of IL-6 was determined in amniotic fluid, cervicovaginal fluid, and maternal blood. In concordance with our review, the association between IL-6 and preterm birth was significant in amniotic fluid and cervicovaginal fluid but not in maternal blood (69). However, further validation and refinement of these tests could enhance their clinical utility and reduce reliance on invasive diagnostic procedures also in PPROM.

The presence of IAI among women with PPROM is associated with a shorter latent period and with lower gestational age at delivery. The gestational age is the main determinant of neonatal outcome (48,56). However, there has been an ongoing debate as to whether IAI itself worsens the neonatal outcomes. The presence of elevated immune proteins associated with IAI can lead to the development of acute histological chorioamnionitis, followed by the activation of the fetal immune system (fetal inflammatory response syndrome), which might progress toward multiorgan dysfunction and failure (70–72). Our study shows that the upregulation of amniotic fluid IL-6 is associated with a higher rate of bronchopulmonary dysplasia, RDS, and EONS. This is in line with similar studies (6,7). Interestingly, we didn’t find any difference between microbial-associated IAI and sterile IAI in the rate of neonatal complications. From a clinical point of view, it is important that early diagnosis and antibiotic treatment can reduce intra-amniotic inflammatory response in PPROM pregnancies complicated by microbial-associated IAI and sterile IAI (34). This can translate into a longer latent period, higher gestational age at delivery and reduced neonatal morbidity.

The presence of MIAC is typically associated with concurrent IAI (microbial-associated IAI). However, an alternative situation, where MIAC is not accompanied by IAI (colonization), has been previously reported among women with PPROM (35,52). The predominant theory regarding the development of MIAC is the ascension of microbes from the lower genital tract into the amniotic cavity (65,66). The bacterial profiles of amniotic fluid in both phenotypes of MIAC support the theory, because they are largely consistent with those of the vagina (73,74). Our study identified genital mycoplasmas (mainly *Ureaplasma* spp.) as the most common pathogens in amniotic fluid of women with PPROM, which is in line with the existing evidence. However, unlike for microbial-associated IAI, the load of Ureaplasma spp. DNA in the amniotic fluid of women with colonization is significantly lower (35). Given the dose-dependent relationship between the intra-amniotic inflammatory response and the amniotic fluid burden of *Ureaplasma* spp. (56,75), it can be deduced that women with *Ureaplasma* spp. colonization may not elicit an intra-amniotic inflammatory response of sufficient intensity to exceed a clinical threshold for IAI. Our study identified other differences between microbial-associated IAI and colonization. The presence of *Ch. trachomatis*, *S. anginosus*, and *F. nucleatum* was significantly associated with microbial-associated IAI, not colonization. *Ch. trachomatis* is a common sexually transmitted bacteria that leads to infections of the human genital tract (76,77). Multiple sources identified maternal genital *Ch. trachomatis* infection as a risk factor for preterm birth (78,79). The presence of *Ch. trachomatis* in women with PPROM was assessed by Kacerovsky et al. (80). Unlike studies included in this review, Kacerovsky et al. found that the presence of *Ch. trachomatis* in amniotic fluid of women with PPROM was not associated with intensive intra-amniotic inflammatory responses nor with adverse short-term neonatal outcome (80). *F. nucleatum* is one of the most abundant species in the oral cavity and is involved in the pathogenesis of periodontal disease (81). *F. nucleatum* was identified in women with preterm birth associated with IAI. *F nucleatum* may translocate from the maternal oral cavity to the intrauterine cavity *via* hematogenous transmission, which probably represents a minor way of MIAC (82,83). It has been demonstrated that *F. nucleatum* can trigger strong placental inflammation through maternal TLR4-mediated signaling (84).

### Strength and limitations

The major strength of this study is that we did a comprehensive search across several databases without limiting the search to amniotic fluid, but we extended our search to support the transfer of IAI diagnostics from transabdominal amniocentesis to the diagnostics using maternal peripheral blood, cervical fluid, and/or vaginal fluid. In addition, we included the studies investigating the concentrations of different IAI markers to assess the versatility of IL-6. The limitation of this study is that only studies written in English were included in this review. Further, significant heterogeneity was seen in our meta-analysis, namely in the stratification of microbial-associated and sterile IAI, the reflection of global proinflammatory response, and the non-invasive model for diagnosis IAI. However, this heterogeneity could not be solved with a subgroup analysis, since there is a great variance in the size of cohorts, IL-6 concentrations depend on the selected analytical method, and the kind of collected samples in the included studies. Finally, we analyzed several variables in the form of proteins, WBC, glucose and PGE2.

### Conclusions and implications

Quantification of IL-6 is easily available, versatile and rapid method to recognize both microbial-associated and sterile IAI in PPROM. IL-6 concentration in amniotic fluid positively correlates with IL-6 concentration in cervical and vaginal fluid and therefore, further research may shift the diagnosis of IAI towards non-invasive and more considerate care of pregnant women. Further, elevated IL-6 concentration is associated with the presence of obligatory pathogens and, as we confirmed, the IL-6 concentration significantly increased if microbial-associated IAI is present in comparison with sterile IAI. This finding enables to stratify the PPROM women with the highest risk of severe consequences caused by the presence of pathogens. On the other hand, there is no difference in the proportion of neonatal morbidity if microbial-associated and sterile IAI are compared.

## Data Availability

All data produced in the present study are available upon reasonable request to the authors

## CRediT authorship contribution statement

**Marie Vajrychova:** conceptualization, literature search, data synthesis, writing – original draft, review & editing. **Michaela Sadibolova:** literature search, data curation, writing – review & editing, **Rudolf Kukla:** data curation, writing – review & editing. **Radka Bolehovska:** data curation, writing – review & editing. **Marian Kacerovsky:** supervision, writing – review & editing. **Jaroslav Stranik:** conceptualization, literature search, writing – original draft, review & editing.

## Acknowledgement

Supported by Ministry of Health of the Czech Republic, grant nr. NU21J-07-00058 and MH CZ-DRO (UHHK, 00179906).

## Reference

1. Blencowe H, Cousens S, Oestergaard MZ, Chou D, Moller AB, Narwal R, et al. National, regional, and worldwide estimates of preterm birth rates in the year 2010 with time trends since 1990 for selected countries: a systematic analysis and implications. Lancet. 2012 Jun 9;379(9832):2162–72.

2. Perin J, Mulick A, Yeung D, Villavicencio F, Lopez G, Strong KL, et al. Global, regional, and national causes of under-5 mortality in 2000-19: an updated systematic analysis with implications for the Sustainable Development Goals. Lancet Child Adolesc Health. 2022 Feb;6(2):106–15.

3. Walani SR. Global burden of preterm birth. Int J Gynaecol Obstet. 2020 Jul;150(1):31–3.

4. Mercer BM. Preterm Premature Rupture of the Membranes. Obstetrics & Gynecology. 2003 Jan;101(1):178.

5. Tsakiridis I, Mamopoulos A, Chalkia-Prapa EM, Athanasiadis A, Dagklis T. Preterm Premature Rupture of Membranes: A Review of 3 National Guidelines. Obstetrical & Gynecological Survey. 2018 Jun;73(6):368.

6. Romero R, Miranda J, Chaemsaithong P, Chaiworapongsa T, Kusanovic JP, Dong Z, et al. Sterile and microbial-associated intra-amniotic inflammation in preterm prelabor rupture of membranes. The Journal of Maternal-Fetal & Neonatal Medicine. 2015 Aug 13;28(12):1394–409.

7. Shim SS, Romero R, Hong JS, Park CW, Jun JK, Il Kim B, et al. Clinical significance of intra-amniotic inflammation in patients with preterm premature rupture of membranes. American Journal of Obstetrics and Gynecology. 2004 Oct 1;191(4):1339–45.

8. Romero R, Gomez-Lopez N, Winters AD, Jung E, Shaman M, Bieda J, et al. Evidence that intra-amniotic infections are often the result of an ascending invasion – a molecular microbiological study. Journal of Perinatal Medicine. 2019 Nov 1;47(9):915–31.

9. Musilova I, Kutová R, Pliskova L, Stepan M, Menon R, Jacobsson B, et al. Intraamniotic Inflammation in Women with Preterm Prelabor Rupture of Membranes. PLOS ONE. 2015 7;10(7):e0133929.

10. Musilova I, Andrys C, Drahosova M, Zednikova B, Hornychova H, Pliskova L, et al. Late preterm prelabor rupture of fetal membranes: fetal inflammatory response and neonatal outcome. Pediatr Res. 2018 Mar;83(3):630–7.

11. Kacerovsky M, Holeckova, Magdalena, Stepan, Martin, Gregor, Miroslav, Vescicik, Peter, Lesko, Daniel, et al. Amniotic fluid glucose level in PPROM pregnancies: a glance at the old friend. The Journal of Maternal-Fetal & Neonatal Medicine. 2022 Jun 18;35(12):2247–59.

12. Harirah H, Donia SE, Hsu CD. Amniotic Fluid Matrix Metalloproteinase-9 and Interleukin-6 in Predicting Intra-Amniotic Infection. Obstetrics & Gynecology. 2002 Jan;99(1):80.

13. Musilova I, Andrys C, Holeckova M, Kolarova V, Pliskova L, Drahosova M, et al. Interleukin-6 measured using the automated electrochemiluminescence immunoassay method for the identification of intra-amniotic inflammation in preterm prelabor rupture of membranes. The Journal of Maternal-Fetal & Neonatal Medicine. 2020 Jun 2;33(11):1919– 26.

14. Romero R, Yoon BH, Mazor M, Gomez R, Diamond MP, Kenney JS, et al. The diagnostic and prognostic value of amniotic fluid white blood cell count, glucose, interleukin-6, and gram stain in patients with preterm labor and intact membranes. Am J Obstet Gynecol. 1993 Oct;169(4):805–16.

15. Chaemsaithong P, Romero R, Korzeniewski SJ, Martinez-Varea A, Dong Z, Yoon BH, et al. A point of care test for interleukin-6 in amniotic fluid in preterm prelabor rupture of membranes: a step toward the early treatment of acute intra-amniotic inflammation/infection. The Journal of Maternal-Fetal & Neonatal Medicine. 2016 Feb 1;29(3):360–7.

16. Kacerovsky M, Musilova I, Bestvina T, Stepan M, Cobo T, Jacobsson B. Preterm Prelabor Rupture of Membranes between 34 and 37 Weeks: A Point-of-Care Test of Vaginal Fluid Interleukin-6 Concentrations for a Noninvasive Detection of Intra-Amniotic Inflammation. Fetal Diagn Ther. 2018;43(3):175–83.

17. Cobo T, Kacerovsky M, Jacobsson B. Noninvasive Sampling of the Intrauterine Environment in Women with Preterm Labor and Intact Membranes. Fetal Diagn Ther. 2018;43(4):241–9.

18. Page MJ, Moher D, Bossuyt PM, Boutron I, Hoffmann TC, Mulrow CD, et al. PRISMA 2020 explanation and elaboration: updated guidance and exemplars for reporting systematic reviews. BMJ. 2021 Mar 29;372:n160.

19. Ouzzani M, Hammady H, Fedorowicz Z, Elmagarmid A. Rayyan—a web and mobile app for systematic reviews. Syst Rev. 2016 Dec;5(1):210.

20. Balduzzi S, Rücker G, Schwarzer G. How to perform a meta-analysis with R: a practical tutorial. Evid Based Ment Health. 2019 Nov;22(4):153–60.

21. Viechtbauer W. Conducting Meta-Analyses in R with the metafor Package. Journal of Statistical Software. 2010 Aug 5;36:1–48.

22. Companion R Package for the Guide Doing Meta-Analysis in R [Internet]. [cited 2025 Mar 29]. Available from: https://dmetar.protectlab.org/

23. McGrath S, Zhao X, Ozturk O, Katzenschlager S, Steele R, Benedetti A. metamedian: An R package for meta-analyzing studies reporting medians. Res Synth Methods. 2024 Mar;15(2):332–46.

24. Wickham H, Averick M, Bryan J, Chang W, McGowan LD, François R, et al. Welcome to the Tidyverse. Journal of Open Source Software. 2019 Nov 21;4(43):1686.

25. Harrer M, Cuijpers P, Furukawa T, Ebert D. Doing Meta-Analysis with R: A Hands-On Guide. New York: Chapman and Hall/CRC; 2021. 500 p.

26. Aberšek N, Tsiartas P, Soucek O, Andrys C, Musilova I, Jacobsson B, et al. Characterizing of intra-amniotic inflammatory changes associated with chronic inflammation in the placenta marked by elevated amniotic fluid interferon gamma-induced protein 10 (IP-10) in pregnancies complicated by preterm prelabor rupture of membranes. Eur J Obstet Gynecol Reprod Biol. 2024 May;296:292–8.

27. Andrys C, Musilova I, Drahosova M, Soucek O, Pliskova L, Jacobsson B, et al. Cervical fluid calreticulin and cathepsin-G in pregnancies complicated by preterm prelabor rupture of membranes. J Matern Fetal Neonatal Med. 2018 Feb;31(4):481–8.

28. Back JH, Kim SY, Gu MB, Kim HJ, Lee KN, Lee JE, et al. Proteomic analysis of plasma to identify novel biomarkers for intra-amniotic infection and/or inflammation in preterm premature rupture of membranes. Sci Rep. 2023 Apr 6;13(1):5658.

29. Fulova V, Hostinska E, Studnickova M, Huml K, Zapletalova J, Halek J, et al. Transabdominal amniocentesis in expectant management of preterm premature rupture of membranes: A single center prospective study. Biomed Pap Med Fac Univ Palacky Olomouc Czech Repub. 2021 Sep;165(3):305–15.

30. Gulati S, Agrawal S, Raghunandan C, Bhattacharya J, Saili A, Agarwal S, et al. Maternal serum interleukin-6 and its association with clinicopathological infectious morbidity in preterm premature rupture of membranes: a prospective cohort study. J Matern Fetal Neonatal Med. 2012 Aug;25(8):1428–32.

31. Ikeda M, Oshima Y, Tsumura K, Gondo K, Ono T, Kozuma Y, et al. Antibiotic administration reduced intra-amniotic inflammation 7 days after preterm premature rupture of the membranes with intra-amniotic infection. The Journal of Maternal-Fetal & Neonatal Medicine. 2023 Dec 15;36(2):2286189.

32. Joo E, Park KH, Kim YM, Ahn K, Hong S. Maternal Plasma and Amniotic Fluid LBP, Pentraxin 3, Resistin, and IGFBP-3: Biomarkers of Microbial Invasion of Amniotic Cavity and/or Intra-amniotic Inflammation in Women with Preterm Premature Rupture of Membranes. J Korean Med Sci. 2021 Nov 15;36(44):e279.

33. Jun JK, Yoon BH, Romero R, Kim M, Moon JB, Ki SH, et al. Interleukin 6 determinations in cervical fluid have diagnostic and prognostic value in preterm premature rupture of membranes. Am J Obstet Gynecol. 2000 Oct;183(4):868–73.

34. Kacerovsky M, Romero R, Stepan M, Stranik J, Maly J, Pliskova L, et al. Antibiotic administration reduces the rate of intraamniotic inflammation in preterm prelabor rupture of the membranes. American Journal of Obstetrics and Gynecology. 2020 Jul 1;223(1):114.e1-114.e20.

35. Kacerovsky M, Stranik J, Matulova J, Chalupska M, Mls J, Faist T, et al. Clinical characteristics of colonization of the amniotic cavity in women with preterm prelabor rupture of membranes, a retrospective study. Sci Rep. 2022 Mar 24;12(1):5062.

36. Kacerovsky M, Kukla R, Bolehovska R, Bostik P, Matulova J, Mls J, et al. Prevalence and Load of Cervical Ureaplasma Species With Respect to Intra-amniotic Complications in Women With Preterm Prelabor Rupture of Membranes Before 34 weeks. Front Pharmacol [Internet]. 2022 Mar 31 [cited 2025 Mar 28];13. Available from: https://www.frontiersin.org/journals/pharmacology/articles/10.3389/fphar.2022.860498/full

37. Lee SM, Park KH, Jung EY, Kook SY, Park H, Jeon SJ. Inflammatory proteins in maternal plasma, cervicovaginal and amniotic fluids as predictors of intra-amniotic infection in preterm premature rupture of membranes. PLOS ONE. 2018 7;13(7):e0200311.

38. Lewis DF, Barrilleaux PS, Wang Y, Adair CD, Baier J, Kruger T. Detection of interleukin-6 in maternal plasma predicts neonatal and infectious complications in preterm premature rupture of membranes. Am J Perinatol. 2001 Nov;18(7):387–91.

39. Musilova I, Andrys C, Drahosova M, Hornychova H, Jacobsson B, Menon R, et al. Amniotic fluid prostaglandin E2 in pregnancies complicated by preterm prelabor rupture of the membranes. J Matern Fetal Neonatal Med. 2016 Sep;29(18):2915–23.

40. Musilova I, Bestvina T, Hudeckova M, Michalec I, Cobo T, Jacobsson B, et al. Vaginal fluid interleukin-6 concentrations as a point-of-care test is of value in women with preterm prelabor rupture of membranes. American Journal of Obstetrics and Gynecology. 2016 Nov 1;215(5):619.e1–619.e12.

41. Musilova I, Andrys C, Drahosova M, Soucek O, Kutova R, Pliskova L, et al. Amniotic fluid calreticulin in pregnancies complicated by the preterm prelabor rupture of membranes. J Matern Fetal Neonatal Med. 2016 Dec;29(24):3921–9.

42. Musilova I, Pliskova L, Gerychova R, Janku P, Simetka O, Matlak P, et al. Maternal white blood cell count cannot identify the presence of microbial invasion of the amniotic cavity or intra-amniotic inflammation in women with preterm prelabor rupture of membranes. PLOS ONE. 2017 12;12(12):e0189394.

43. Musilova I, Kacerovsky M, Stepan M, Bestvina T, Pliskova L, Zednikova B, et al. Maternal serum C-reactive protein concentration and intra-amniotic inflammation in women with preterm prelabor rupture of membranes. PLOS ONE. 2017 8;12(8):e0182731.

44. Musilova I, Andrys C, Drahosova M, Soucek O, Pliskova L, Stepan M, et al. Amniotic fluid clusterin in pregnancies complicated by the preterm prelabor rupture of membranes. J Matern Fetal Neonatal Med. 2017 Nov;30(21):2529–37.

45. Musilova I, Andrys C, Drahosova M, Soucek O, Pliskova L, Stepan M, et al. Amniotic fluid cathepsin-G in pregnancies complicated by the preterm prelabor rupture of membranes. J Matern Fetal Neonatal Med. 2017 Sep;30(17):2097–104.

46. Park KH, Kim SN, Oh KJ, Lee SY, Jeong EH, Ryu A. Noninvasive prediction of intra-amniotic infection and/or inflammation in preterm premature rupture of membranes. Reprod Sci. 2012 Jun;19(6):658–65.

47. Rizzo G, Capponi A, Vlachopoulou A, Angelini E, Grassi C, Romanini C. Interleukin-6 concentrations in cervical secretions in the prediction of intrauterine infection in preterm premature rupture of the membranes. Gynecol Obstet Invest. 1998 Aug;46(2):91–5.

48. Rodríguez-Trujillo A, Cobo T, Vives I, Bosch J, Kacerovsky M, Posadas DE, et al. Gestational age is more important for short-term neonatal outcome than microbial invasion of the amniotic cavity or intra-amniotic inflammation in preterm prelabor rupture of membranes. Acta Obstetricia et Gynecologica Scandinavica. 2016;95(8):926–33.

49. Soucek O, Kacerovsky M, Kacerovska Musilova I, Stranik J, Kukla R, Bolehovska R, et al. Amniotic fluid CD36 in pregnancies complicated by spontaneous preterm delivery: a retrospective cohort study. J Matern Fetal Neonatal Med. 2023 Dec;36(1):2214838.

50. Spacek R, Kacerovský M, Andrýs C, Souček O, Kukla R, Bolehovská R, et al. Amniotic fluid soluble CD93 is elevated in the presence of intra-amniotic inflammation in preterm prelabor rupture of the fetal membranes. Ceska Gynekol. 2022;87(6):388–95.

51. Spacek R, Musilova I, Andrys C, Soucek O, Burckova H, Pavlicek J, et al. Extracellular granzyme A in amniotic fluid is elevated in the presence of sterile intra-amniotic inflammation in preterm prelabor rupture of membranes. J Matern Fetal Neonatal Med. 2022 Sep;35(17):3244–53.

52. Stranik J, Kacerovsky M, Soucek O, Kolackova M, Musilova I, Pliskova L, et al. IgGFc-binding protein in pregnancies complicated by spontaneous preterm delivery: a retrospective cohort study. Sci Rep. 2021 Mar 17;11(1):6107.

53. Theis KR, Romero R, Motomura K, Galaz J, Winters AD, Pacora P, et al. Microbial burden and inflammasome activation in amniotic fluid of patients with preterm prelabor rupture of membranes. J Perinat Med. 2020 Feb 25;48(2):115–31.

54. Musilova I, Andrys C, Drahosova M, Soucek O, Pliskova L, Jacobsson B, et al. Cervical fluid interleukin 6 and intra-amniotic complications of preterm prelabor rupture of membranes. J Matern Fetal Neonatal Med. 2018 Apr;31(7):827–36.

55. Maymon E, Romero R, Chaiworapongsa T, Kim JC, Berman S, Gomez R, et al. Value of amniotic fluid neutrophil collagenase concentrations in preterm premature rupture of membranes. Am J Obstet Gynecol. 2001 Nov;185(5):1143–8.

56. Musilova I, Kutová R, Pliskova L, Stepan M, Menon R, Jacobsson B, et al. Intraamniotic Inflammation in Women with Preterm Prelabor Rupture of Membranes. PLoS One. 2015 Jul 24;10(7):e0133929.

57. Yoon BH, Romero R, Moon JB, Shim SS, Kim M, Kim G, et al. Clinical significance of intra-amniotic inflammation in patients with preterm labor and intact membranes. Am J Obstet Gynecol. 2001 Nov;185(5):1130–6.

58. Fortunato SJ, Menon R. Distinct molecular events suggest different pathways for preterm labor and premature rupture of membranes. American Journal of Obstetrics and Gynecology. 2001 Jun 1;184(7):1399–406.

59. Menon R, Behnia F, Polettini J, Richardson LS. Novel pathways of inflammation in human fetal membranes associated with preterm birth and preterm pre-labor rupture of the membranes. Semin Immunopathol. 2020 Aug 1;42(4):431–50.

60. Svenvik M, Jenmalm MC, Brudin L, Raffetseder J, Hellberg S, Axelsson D, et al. Chemokine and cytokine profiles in preterm and term labor, in preterm prelabor rupture of the membranes, and in normal pregnancy. Journal of Reproductive Immunology. 2024 Aug 1;164:104278.

61. Romero R, Miranda J, Chaiworapongsa T, Chaemsaithong P, Gotsch F, Dong Z, et al. A Novel Molecular Microbiologic Technique for the Rapid Diagnosis of Microbial Invasion of the Amniotic Cavity and Intra-Amniotic Infection in Preterm Labor with Intact Membranes. American Journal of Reproductive Immunology. 2014;71(4):330–58.

62. Jalava J, Mäntymaa ML, Ekblad U, Toivanen P, Skurnik M, Lassila O, et al. Bacterial 16S rDNA polymerase chain reaction in the detection of intra-amniotic infection. BJOG: An International Journal of Obstetrics & Gynaecology. 1996;103(7):664–9.

63. Silver RM, Schwinzer B, McGregor JA. Interleukin-6 levels in amniotic fluid in normal and abnormal pregnancies: Preeclampsia, small-for-gestational-age fetus, and premature labor. American Journal of Obstetrics and Gynecology. 1993 Nov 1;169(5):1101– 5.

64. Andrews W, Hauth J, Goldenberg R, Gomez R, Romero R, Cassell G. Amniotic fluid interleukin-6: correlation with upper genital tract microbial colonization and gestational age in women delivered after spontaneous labor versus indicated delivery. American journal of obstetrics and gynecology [Internet]. 1995 Aug [cited 2025 Apr 18];173(2). Available from: https://pubmed.ncbi.nlm.nih.gov/7645642/

65. Gomez-Lopez N, Galaz J, Miller D, Farias-Jofre M, Liu Z, Arenas-Hernandez M, et al. The immunobiology of preterm labor and birth: intra-amniotic inflammation or breakdown of maternal–fetal homeostasis. 2022 Aug 1 [cited 2025 Apr 19]; Available from: https://rep.bioscientifica.com/view/journals/rep/164/2/REP-22-0046.xml

66. Romero R, Espinoza J, Gonçalves LF, Kusanovic JP, Friel L, Hassan S. The Role of Inflammation and Infection in Preterm Birth. Seminars in Reproductive Medicine. 2007 Jan 5;25:21–39.

67. Musilova I, Bestvina T, Stranik J, Stepan M, Jacobsson B, Kacerovsky M. Transabdominal Amniocentesis Is a Feasible and Safe Procedure in Preterm Prelabor Rupture of Membranes. Fetal Diagnosis and Therapy. 2017 Feb 25;42(4):257–61.

68. Kacerovsky M, Musilova I, Hornychova H, Kutova R, Pliskova L, Kostal M, et al. Bedside assessment of amniotic fluid interleukin-6 in preterm prelabor rupture of membranes. American Journal of Obstetrics and Gynecology. 2014 Oct 1;211(4):385.e1–385.e9.

69. Chang Y, Li W, Shen Y, Li S, Chen X. Association between interleukin-6 and preterm birth: a meta-analysis. Ann Med. 55(2):2284384.

70. Kim CJ, Romero R, Chaemsaithong P, Chaiyasit N, Yoon BH, Kim YM. Acute chorioamnionitis and funisitis: definition, pathologic features, and clinical significance. Am J Obstet Gynecol. 2015 Oct;213(4 Suppl):S29–52.

71. Hofer N, Kothari R, Morris N, Müller W, Resch B. The fetal inflammatory response syndrome is a risk factor for morbidity in preterm neonates. American Journal of Obstetrics and Gynecology. 2013 Dec 1;209(6):542.e1–542.e11.

72. Romero R, Savasan ZA, Chaiworapongsa T, Berry SM, Kusanovic JP, Hassan SS, et al. Hematologic profile of the fetus with systemic inflammatory response syndrome. Journal of Perinatal Medicine. 2012 Jan 1;40(1):19–32.

73. Romero R, Gomez-Lopez N, Winters AD, Jung E, Shaman M, Bieda J, et al. Evidence that intra-amniotic infections are often the result of an ascending invasion - a molecular microbiological study. J Perinat Med. 2019 Nov 26;47(9):915–31.

74. DiGiulio DB, Romero R, Kusanovic JP, Gómez R, Kim CJ, Seok KS, et al. Prevalence and diversity of microbes in the amniotic fluid, the fetal inflammatory response, and pregnancy outcome in women with preterm pre-labor rupture of membranes. Am J Reprod Immunol. 2010 Jul 1;64(1):38–57.

75. Jacobsson B, Aaltonen R, Rantakokko-Jalava K, Morken NH, Alanen A. Quantification of Ureaplasma urealyticum DNA in the amniotic fluid from patients in PTL and pPROM and its relation to inflammatory cytokine levels. Acta Obstet Gynecol Scand. 2009;88(1):63–70.

76. Unemo M, Bradshaw CS, Hocking JS, Vries HJC de, Francis SC, Mabey D, et al. Sexually transmitted infections: challenges ahead. The Lancet Infectious Diseases. 2017 Aug 1;17(8):e235–79.

77. Tuddenham S, Hamill MM, Ghanem KG. Diagnosis and Treatment of Sexually Transmitted Infections: A Review. JAMA. 2022 Jan 11;327(2):161–72.

78. Andrews WW, Goldenberg RL, Mercer B, Iams J, Meis P, Moawad A, et al. The Preterm Prediction Study: Association of second-trimester genitourinary chlamydia infection with subsequent spontaneous preterm birth. American Journal of Obstetrics and Gynecology. 2000 Sep 1;183(3):662–8.

79. Ahmadi A, Ramazanzadeh R, Sayehmiri K, Sayehmiri F, Amirmozafari N. Association of Chlamydia trachomatis infections with preterm delivery; a systematic review and meta-analysis. BMC Pregnancy and Childbirth. 2018 Jun 18;18(1):240.

80. Kacerovsky M, Romero R, Pliskova L, Bolehovska R, Hornychova H, Matejkova A, et al. Presence of Chlamydia trachomatis DNA in the amniotic fluid in women with preterm prelabor rupture of membranes. J Matern Fetal Neonatal Med. 2021 May;34(10):1586–97.

81. Han YW. Fusobacterium nucleatum: a commensal-turned pathogen. Curr Opin Microbiol. 2015 Feb;0:141–7.

82. Radochova V, Stepan M, Kacerovska Musilova I, Slezak R, Vescicik P, Menon R, et al. Association between periodontal disease and preterm prelabour rupture of membranes. Journal of Clinical Periodontology. 2019;46(2):189–96.

83. Radochova V, Kacerovska Musilova, Ivana, Stepan, Martin, Vescicik, Peter, Slezak, Radovan, Jacobsson, Bo, et al. Periodontal disease and intra-amniotic complications in women with preterm prelabor rupture of membranes. The Journal of Maternal-Fetal & Neonatal Medicine. 2018 Nov 2;31(21):2852–61.

84. Ghosh A, Jaaback K, Boulton A, Wong-Brown M, Raymond S, Dutta P, et al. Fusobacterium nucleatum: An Overview of Evidence, Demi-Decadal Trends, and Its Role in Adverse Pregnancy Outcomes and Various Gynecological Diseases, including Cancers. Cells. 2024 Jan;13(8):717.

